# Small-Sized Reasoning Language Models for Linguistic Screening of Alzheimer’s Disease

**DOI:** 10.64898/2025.12.24.25342972

**Authors:** Venkatanand R. Addepalli, Nader Abdalnabi, Erich Kummerfeld, Guy Hembroff, Andrew Kiselica, Praveen Rao, Knoo Lee

**Affiliations:** Institute for Data Science and Informatics, University of Missouri, Columbia, Missouri, USA; Institute of Health Informatics, University of Minnesota, Twin Cities, Minnesota, USA; Applied Computing, Michigan Technological University, Houghton, Michigan, USA; Institute of Gerontology, Health Policy & Management, University of Georgia, Athens, Georgia, USA; Electrical Engineering and Computer Science, University of Missouri, Columbia, Missouri, USA; Sinclair School of Nursing, University of Missouri, Columbia, Missouri, USA

## Abstract

Alzheimer’s disease (AD) is increasing in prevalence, and early detection is essential for timely care. Clinical services face growing demand, leading to delays in diagnostic appointments and increasing the risk of disease progression before evaluation. This work examines artificial intelligence (AI) methods for assessing cognitive status from linguistic features. The proposed architecture uses small language models (SLMs) to analyze speech patterns, and its compact design allows deployment on mobile devices. Recent reasoning-focused models, including Deepseek-R1 and Llama, were evaluated for dementia classification. Multiple fine-tuning strategies were compared, and the best model achieved 91% accuracy and an F1 score. The findings show that AI systems built on SLMs can achieve performance comparable to large language models, indicating their potential as efficient tools that may support health care providers through accessible pre-clinical screening for AD.

## Introduction

Dementia is a rapidly escalating neurodegenerative disorder known to disrupt daily functioning and independence in older adults^1^. In the United States, approximately 7.2 million individuals aged 65 years and older are living with the disease, and more than 70 % of these cases occur in persons aged 75 years or older. Clinical experts also predict that these numbers can be doubled by 2060^2^.

The escalating prevalence of dementia highlights the urgent need for more effective screening strategies and precise diagnostic tools^3^. Screening, however, requires continuous observation over extended periods, a logistical challenge that limits healthcare providers’ ability to monitor every individual at risk^4^. Neuroimaging modalities - including amyloidPET and structural MRI ^5^-have the capacity to detect characteristic brain alterations, yet their routine use is constrained by radiation exposure, limited accessibility, and high operational costs^6^. While cerebrospinal fluid (CSF) assays for phosphorylated tau offer a sensitive biomarker of Alzheimer’s disease progression, lumbar puncture remains invasive and is frequently declined by patients^7^. Recent studies have identified plasma amyloid β and tau as promising blood-based biomarkers^8–10^. In contrast, retinal imaging of amyloid-β and tau deposits, salivary proteomics, and digital behavioral metrics (speech patterns, gait dynamics) have also shown potential for early risk stratification^11^. Among clinicians, cognitive biomarkers remain the most widely used, primarily because they are simple to administer and supported by validated instruments, such as the Mini-Mental State Examination (MMSE) and the Montreal Cognitive Assessment (MoCA)^12^. Speech-language analysis may offer a low-cost, non-invasive alternative that can be integrated into routine care, allowing patients who suspect early cognitive decline to obtain timely evaluation and guidance^13^.

A study ^14^ highlighted that specific linguistic traits can aid primary-care clinicians in gauging disease progression and estimating the stage of Alzheimer’s disease and related dementia (ADRD)^15^. When combined with the results of standardized cognitive instruments^16^, these speech-based markers show promise in offering a complementary route for clinicians to assess a patient’s performance across various contexts, including memory recall, verbal fluency, and descriptive narration.

A current trend in data science and clinical practice is the use of biomarkers derived from unstructured data across multiple modalities, supported by extensive clinical datasets ^17–19^. Machine learning (ML) has become common in ADRD research, spanning MRI, PET, clinical records, and genomic data ^20^. Traditional ML methods remain limited by reliance on manual feature engineering and poor scalability to complex inputs ^21^. Deep learning models achieve strong diagnostic performance ^22–25^, but their opaque decision processes hinder clinical acceptance. Natural language processing approaches have also been applied to patient-generated text, though their dependence on predefined features restricts flexibility and explainability ^26^. These challenges, together with the growth of large-language models (LLMs) capabilities, motivated our development of a more interpretable framework for linguistic biomarker analysis in ADRD^27^.

Yet the majority of cutting-edge LLMs are delivered exclusively via cloud-based services, creating a set of practical and regulatory challenges for healthcare institutions^28^. Transmitting protected health information (PHI) to remote APIs risks non-compliance with privacy laws and exposes systems to data breach threats and the collateral hazards of malware or ransomware attacks. Moreover, long-term reliance on externally maintained models can introduce undocumented behavior, integration friction, and maintenance uncertainty, thereby elevating institutional liability^29^. In addition to these security concerns, cloud-hosted LLMs often suffer from sporadic availability, a problem that has manifested in notable service outages across major cloud platforms^30^. These reliability issues, coupled with the high computational and energy requirements of real-time inference, hinder the practical deployment of LLMs in frontline clinical workflows where instant, on-premises analysis of notes, discharge summaries, or triage reports is essential^30^. To address the privacy and reliability limitations of cloud-based LLMs, we propose using small language models (SLMs) with fewer than 2 billion parameters. Unlike LLMs, these models can run entirely on local hardware, thereby preventing the exposure of protected health information and supporting compliance with stringent data governance requirements. Recent findings suggest that SLMs can perform comparably to much larger models while requiring far less computational power and energy^31^. This efficiency makes them feasible for clinics with limited resources and for home-based screening, offering a practical and private approach to identifying early signs of ADRD. Their ability to run on commodity hardware enables broad, scalable deployment in healthcare settings.

## Methods

### Data source

The present study utilized the Pitt Corpus, a publicly available resource housed in the DementiaBank database^32^. The corpus is organized into four distinct subsections: Boston Cookie Theft picture description, Fluency, Recall, and Sentence Construction. Each section comprises synchronized audio recordings and their corresponding transcriptions from three participant groups: individuals diagnosed with dementia, those with mild cognitive impairment (MCI), and healthy controls. The subsection we focus on is adapted from the Boston Aphasia Exam protocol^33^, in which a participant and an examiner occupy a quiet room. The examiner presents a standardized picture (the “Cookie Theft” scene, see Figure 1), and the participant is prompted to describe what they observe verbally. The raw recordings were transcribed into textual form using the CLAN (Computerized Language Analysis) software suite^34^, a tool widely employed for the precise annotation of speech corpora.

**Figure 1.**
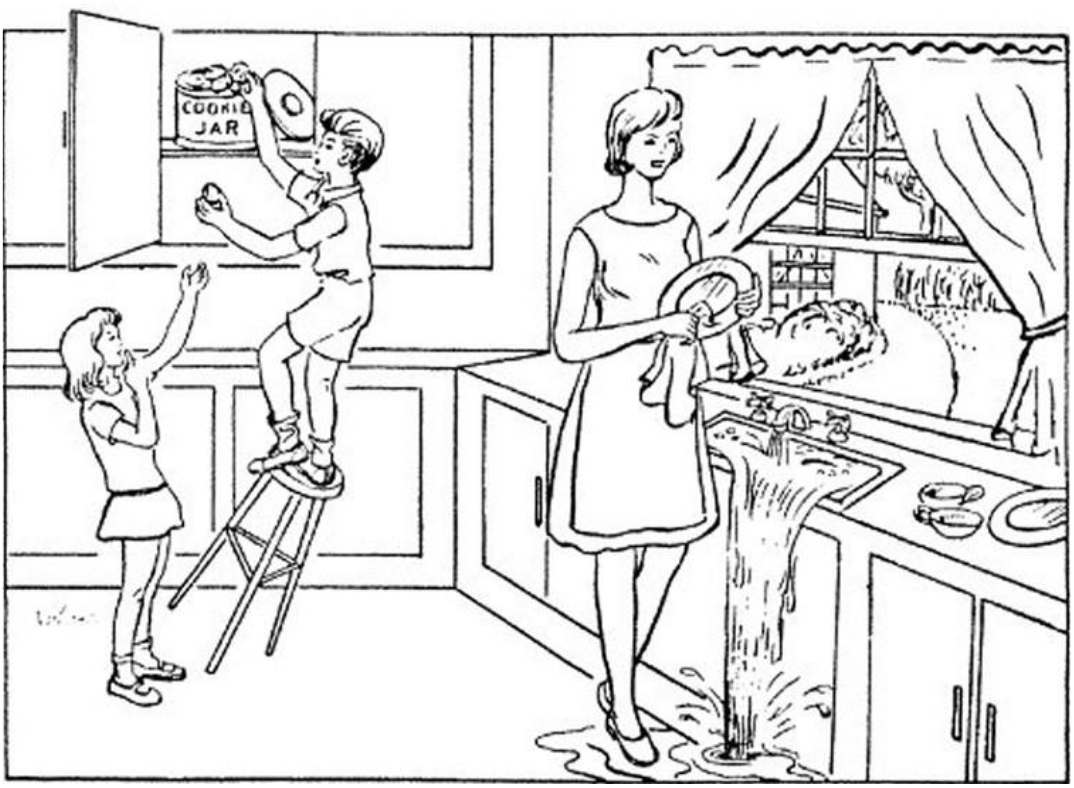
Boston cookie theft picture^35^

In this study, we obtained the transcripts from the Cookie Theft picture description task included in the Pitt Corpus of DementiaBank^35^. We analyzed 549 recordings from 268 unique participants: 243 from healthy controls and 306 from participants with probable AD. Data from other sections of the corpus were not included because the number of control samples was insufficient, and applying data augmentation in this context would likely introduce bias toward the limited number of available healthy control recordings.

### Data Preprocessing

During the analysis, we manually inspected the dataset for diagnosis labels, which included probable AD, possible AD, mild cognitive impairment (MCI), and control. Probable AD and possible AD differ in MMSE scores, with possible AD having a score greater than 20. For our study, probable AD (MMSE < 20) and healthy control samples (MMSE >= 25) were included. This resulted in a total of n=549 total recordings (306 probable AD, 243 controls).

The raw data from the corpus consists of transcripts of the interviewers’ (INV) questions and participants’ (PAR) responses. We used the participants’ responses as input variables, with the corresponding class labels as output variables. Data cleaning involved removing special tokens that are not part of the sentence structure. We preserved filler words (e.g., “uhh”, “umm”) within their respective sentences; all other extraneous content (e.g., [* s:r], [//]) was discarded. This preprocessing procedure was applied uniformly across the entire dataset, encompassing both dementia and control sections of the Cookie Theft task. Finally, the cleaned dataset was split into training (80%), validation (10%), and test (10%) subsets (Table 1 and Figure 2).

**Table 1.**
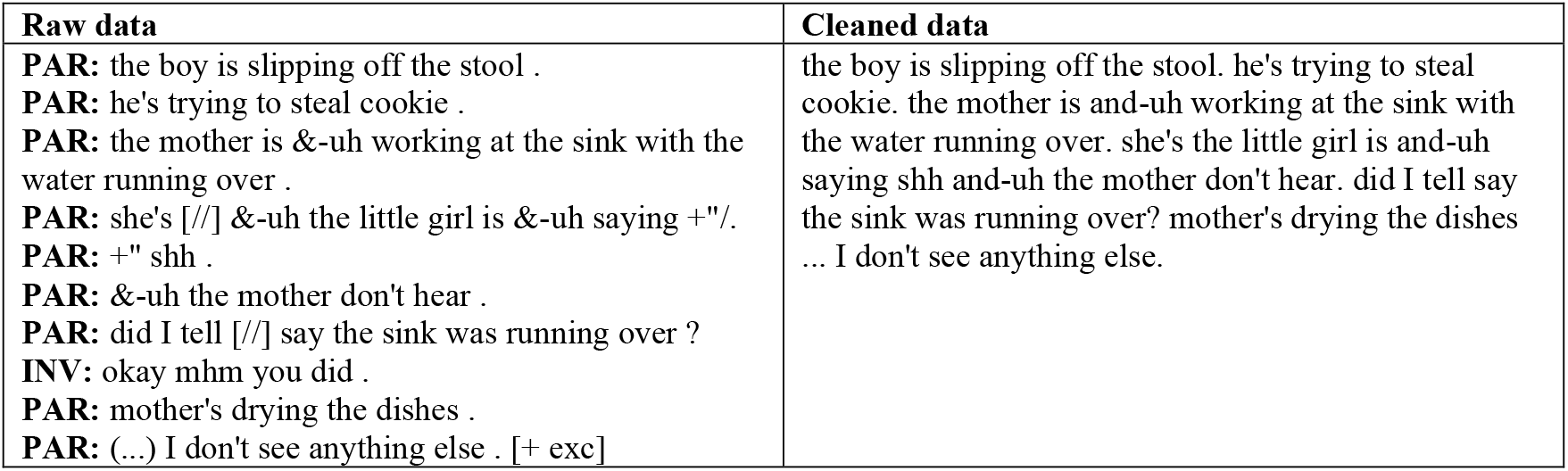
Illustration of the data cleaning process. The table displays paired examples of raw text (left) and the corresponding cleaned text (right), prepared for analysis.

**Figure 2.**
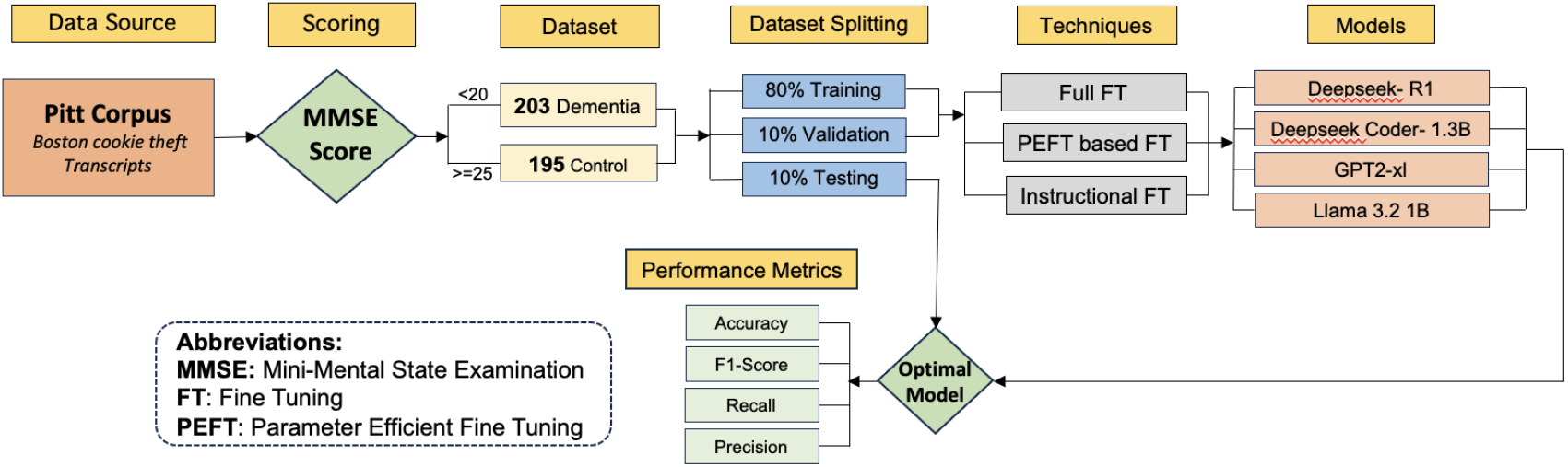
Overview of the Data Analysis Approach.

### Pre-Trained Models and Hyperparameter Optimization

Several studies have explored the application of small language models (SLMs) to text classification problems^31,36,37^. Building on this evidence, we chose to fine-tune four small language models, namely DeepSeek-R1^38^, DeepSeek Coder^39^, OpenAI GPT^40^, and LLaMA 3.2^41^, on the Pitt Corpus dataset. These models belong to the “small” tier of the LLM family and were selected because decoder-based models, initially designed for next-word prediction, can be effectively fine-tuned for classification tasks. Their advanced reasoning capabilities may provide an advantage in classification tasks that require understanding complex contexts or logical relationships in the data (Table 2).

**Table 2.** Comparison of SLMs based on architectural design, parameter size, training data, and functional strengths.

| Model | Architecture | Parameter* | Window | Training Data | Key Highlights | Strengths |
| --- | --- | --- | --- | --- | --- | --- |
| Deepseek-R1 | Decoder-only (mixture-of-experts) | 1.5 B | 16 K | English, Chinese, mixed-domain text | RL-based fine-tuning + COT loop | Reasoning, general NLP |
| Deepseek-Coder | Same as above | 1.3 B | 16 K | Code corpora + natural-lang | Code-centric training + distilled variants | Code-generation, multilingual |
| Llama 3.2 | Decoder-only | 1 B | 128 K | 405 B tokens, multi-lang + multimodal | Efficient scaling, large context window | Code-generation, prompt-based reasoning |
| GPT-2 | Decoder-only | 1.5 B | 1 K | English-only (OpenAI dataset) | Zero-shot / few-shot proficiency | General NLP, GLUE |

\*Parameter sizes are reported in billions (B) and context windows in tokens (K).
RL = Reinforcement Learning; COT = Chain-of-Thought; GLUE = General Language Understanding Evaluation. Values and descriptions are approximate and simplified for comparative purposes.

Figure 2 shows the overall process of our data analysis, including the selected models that meet the criteria for SLM. Next, we briefly describe each model. Deepseek-R1 builds on the Deepseek-R1-Zero foundation by integrating reinforcement learning, a mixture-of-experts architecture, and a chain-of-thought (COT) prompting loop. The result is a reasoning engine comparable to ChatGPT-O1, yet its distilled forms (1.5 B, 7 B, 8 B) keep the network compact. Deepseek-Coder (1.3–33 B parameters) surpasses proprietary code models while still retaining substantial English and Chinese language knowledge; we use this as a non-COT baseline. Llama 3.2 1B was trained on a staggering 405 B tokens and can hold 128 K tokens in its context window, along with reasoning capabilities giving it an edge on code-generation and human-evaluation tasks while remaining efficient on modest hardware. Lastly, the classic GPT-2 1.5B, though limited to roughly 1k tokens, still outshines most small NLP models and shows solid zero-shot/few-shot GLUE performance, proving that a lean, decoder-only design can still deliver broad generalizability. Together, these four models provide a concise narrative of how reasoning, code expertise, multilingualism, and computational efficiency are being balanced in current LLM research.

Regarding fine-tuning, we tested three fine-tuning approaches. Full Fine-Tuning (FFT) updated all weights on the Pitt corpus of dementia speech. To reduce computational time, we also used Parameter-Efficient Fine-Tuning (PEFT) with Low-Rank Adaptation (LoRA), freezing most parameters and learning only small adapters. Finally, with a local ChatGPT-OSS model generated Alpaca-style prompts to highlight dementia-specific patterns, the reasoning-based models were fine-tuned on just ten samples in a few-shot setting, yielding functional classifications and insights into the decision logic that help in evaluating the chain-of-thought architectures of the Deepseek R1 and Llama 3,2 models.

### Inference and model usage

After fine-tuning all the models, they were saved locally. For evaluation, we loaded the fine-tuned models and tested them on a CPU. Different evaluation metrics were then calculated using the test dataset. For the reasoning part, we selected a few cases of dementia and healthy controls and tested them on the IFT-based models.

All models were fine-tuned on an NVIDIA A4000 GPU, running CUDA 12.4 with Python 3.12 on Windows. We downloaded the four pre-trained checkpoints from Hugging Face to local storage and set up the accompanying.env files for the remaining dependencies. To identify the most effective hyperparameter combinations (Table 3), we utilized Weights & Biases (Wandb).

**Table 3.** Hyperparameter sweep configuration table.

| Parameter | Type | Range/Values | Goal/Method |
| --- | --- | --- | --- |
| Sweep Method | - | - | Bayes |
| Metric | - | eval loss | Minimize( goal ) |
| learning rate | log uniform values | 3e-6 | 3e-4 |
| batch size | Categorical | [4, 8, 16, 32] | - |
| epochs | Categorical | [5, 10, 15] | - |
| warmup_ratio | Categorical | [0.01, 0.1] | - |
| weight_decay | Categorical | [0.01, 0.1] | - |

## Results

After updating all the weights, we evaluated each model on the same test set containing 60 samples. DeepSeekR1 emerged as the strongest performer, achieving an Accuracy and F1 score of 91% (Figure 3). Its training/fine-tuning time, however, was longer than that of the other models, taking 1.2 hours for each set of hyperparameters. By contrast, Llama 3.2 delivered the lowest accuracy and required 40 minutes of training. DeepSeek Coder, despite being initially trained on coding datasets, outperformed both GPT-2 and Llama models.

**Figure 3.**
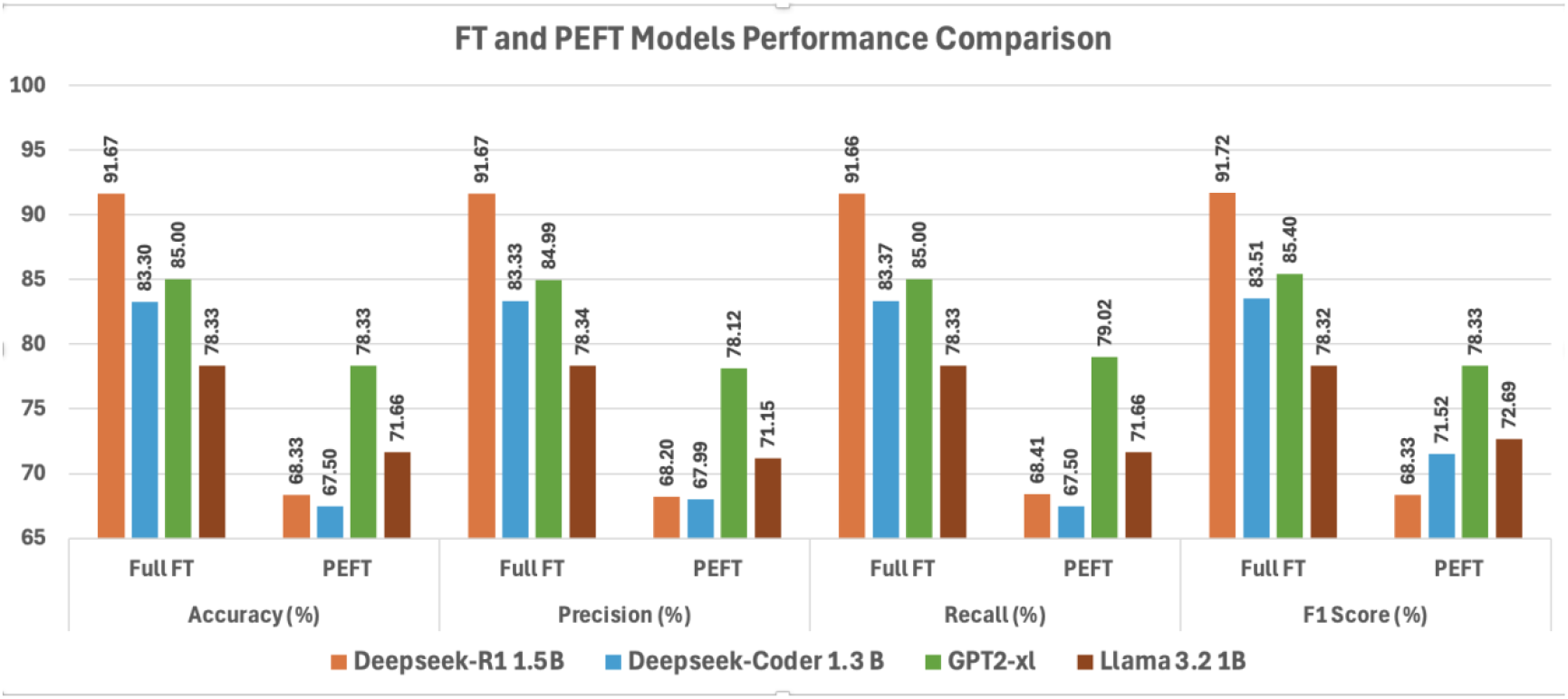
Evaluation across models in different fine-tuned models Full Fine-Tuned Models Comparison

### Fine-Tuned Models Comparis

The validation graphs (Figure 4) showed a steady decline at the beginning, but the validation loss increased beyond 20 epochs. This feature has been observed across all the models in the FFT. For smooth training, we chose to set epcohs between 5 and 15 and logging steps of 100 to 200.

**Figure 4:**
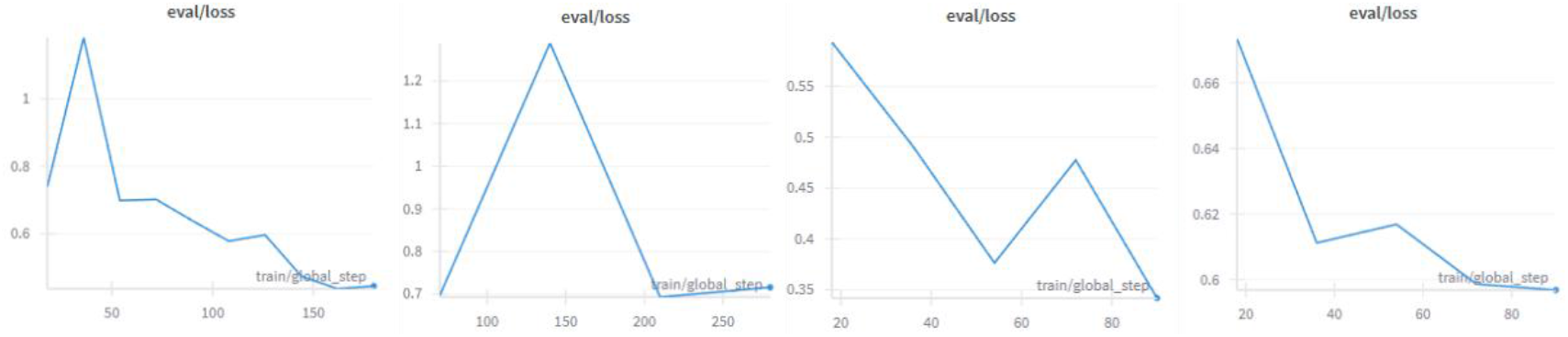
Validation losses for different losses (Deepseek R1, Deepseek Coder, GPT-2 and Llama) PEFT Models Comparison

### PEFT Models Comparison

After pruning the target layers, the models retained less than 1% of their original trainable parameters. This reduction shrank the overall model’s size to nearly 0.28 % and, in most cases, accelerated training time by 93%.

When evaluated, GPT-2 topped the leaderboard with an accuracy of 78.33 % (Figure 3). Surprisingly, DeepSeek R1, despite its Mixture of Experts (MoE) architecture, achieved only 68.33%. The Llama model outperformed the DeepSeek variants, aligning with the expected behavior: Llama was pre-trained on broad language understanding and generation, whereas the other models had been exposed to more math- and coding-specific data.

### Across-Models Comparison

When we fine-tuned the entire network, the models consistently outperformed the PEFT variants in accuracy, f1 score, and recall (Figure 3). On the other hand, the loss curves for the PEFT approach were noticeably smoother, suggesting a steadier learning trajectory.

In general, GPT-2 outperformed Llama, achieving higher test scores in both full-fine-tuning and PEFT setups despite both being decoder-only architectures. The loss curves tell a similar story: Llama models exhibit a gradual, steady decline in validation and training loss, whereas GPT-2’s loss drops sharply, suggesting a more abrupt learning phase. DeepSeek Coder and DeepSeek R1 performed similarly under PEFT, but R1 edged out Coder in classification tasks in both approaches. Overall, the DeepSeek variants excelled at distinguishing between the two classes in our dataset.

### Results from IFT and reasoning outputs

This method incorporates the reasoning abilities of the DeepSeek R1 and Llama 3.2 in classifying dementia/control based on fluency, coherence, information content, syntax, word-finding, repetition, confabulation, and self-correction (Figure 5).

**Figure 5.**
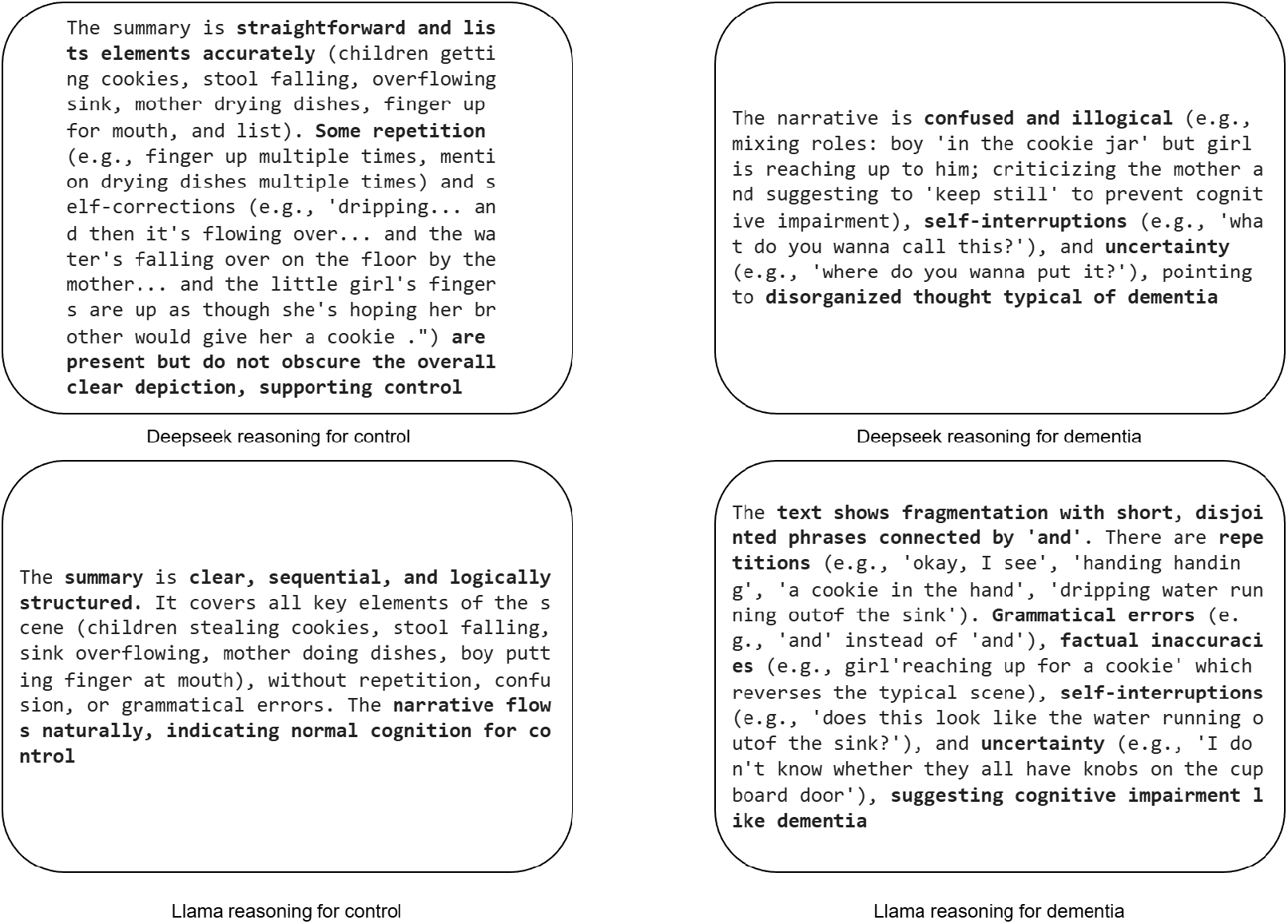
Comparison of reasoning outputs from fine-tuned Deeseek and Llama models

## Discussion

In this study, we fine-tuned a set of SLMs on the Cookie Theft picture-description subset of the Pitt Corpus. This dataset includes a balanced mix of dementia and control transcripts and gives enough samples for reliable testing. All training and inference ran on local CPU or GPU machines, so no cloud access was needed, and patient information stayed on site. We tested three fine-tuning methods, FFT, PEFT, and IFT, and evaluated the models with standard classification metrics and their reasoning outputs.

The best model, DeepSeek R1 1.5B, reached 91 percent accuracy and outperformed previously reported LLMs on the same task while using far fewer resources (Table 4). LLMs need high-power GPUs or TPUs and use much more energy. In contrast, SLMs run on standard hardware, use low energy, and generate minimal heat, which helps clinics with limited budgets or unreliable power. When we compared our fine-tuned SLMs with LLMs on the same data, the SLMs scored higher on every metric while using far less compute. And because the SLMs run locally, no patient data leaves the institution, avoiding the privacy risks seen with cloud-based tools like ChatGPT or DeepSeek 605B.

**Table 4.** Results from the previously fine-tuned LLMs.

| LLMs | Accuracy (%) | F1 Score (%) | Recall | Precision |
| --- | --- | --- | --- | --- |
| GPT-3.5<sup>42</sup> | 58 | 68 |  |  |
| GPT-4.0<sup>42</sup> | 64 | 72 |  |  |
| GPT-4o<sup>42</sup> | 65 | 69 |  |  |
| Llama2 7B<sup>43</sup> | 81.31 | 81.13 | 79.63 | 82.69 |
| BERT<sup>43</sup> | 76.85 | 66.67 | 74.23 | 83.72 |
| Multimodal model (LSTM + Word2Vec)<sup>44</sup> | 69.1 | 62.76 | 62.99 | 64.84 |
| CLIP<sup>45</sup> | 71.97 | 72.00 | 72.02 | 72.05 |
| SLM(Deepseek-R1) | 91.67 | 91.71 | 91.66 | 91.67 |

### Data Privacy and Regulatory Compliance

Although LLMs have demonstrated strong performance for text and speech analysis, they also present several limitations, including risks related to data breaches and security vulnerabilities^46–48^. A prior study^49^ examined these issues by introducing a small amount of medical misinformation (0.001%) into the training data, which resulted in a 4.8% increase in harmful outputs compared with the initial model communications. To address this, we conducted the study using an on-premise SLM deployment, which addresses data privacy and leakage issues. Because all model parameters, training data, and inference outputs remain on the site (e.g., a PC or Raspberry Pi if the model were to be deployed), there is no risk of data leakage. Additionally, we created a pipeline that uses only deidentified data to avoid data leaks and privacy breaches.^36^ Cloud-based LLMs currently lack data transparency, as the data used during pre-training is not fully disclosed, rendering these models essentially as black boxes^50^. Although some LLMs now offer fine-tuning capabilities, many health care providers remain hesitant to rely on them due to their opacity and the uncertainty regarding adherence to medical standards and regulatory requirements^51^. This on-premises deployment of open-source models offers a suitable solution for meeting the requirements of the Health Insurance Portability and Accountability Act (HIPAA) and other regulatory frameworks governing the handling of protected health information.

### Interpretability and Clinical Decision Support

Beyond raw classification performance, the SLMs produced structured reasoning traces that articulated the linguistic cues underlying a dementia prediction. While clinicians did not formally evaluate these explanations, they illustrate the potential of language-model reasoning to aid shared decision-making and to provide clinicians with transparent, interpretable justifications for automated assessments^35^. Stable reasoning traces allow clinicians to notice model errors and build trust through consistent patterns. These cues can guide targeted follow-up questions during remote assessments, making telehealth interactions more efficient^52^. The structured outputs can be logged in electronic health record systems, creating a reviewable trail like other clinical notes^51^.

### Future work

The present study is limited to the cookie theft subset of the Pitt Corpus for both text data and evaluation. Future work should expand this scope to larger, more diverse corpora that include both speech and text, thereby enabling more generalizable findings. Integrating this framework into telemedicine platforms represents a promising direction to assess how different explanation formats influence clinician workload and diagnostic confidence during virtual visits. Reinforcement learning from human feedback may be particularly valuable for enhancing models’ reasoning capabilities, enabling more profound, more clinically relevant insights. Collaboration with linguistic and geriatric experts will be essential for evaluating and refining model outputs using real-time data, allowing iterative adjustments to model parameters and training procedures to improve performance. Further research should also explore how compact reasoning-oriented language models can be incorporated into telemedicine systems as components of modern machine-learning operations (MLOps) pipelines in clinical environments, ensuring reliability, scalability, and seamless clinical integration.

## Conclusion

This study shows that SLMs fine-tuned on a clinically relevant corpus can match dementia-screening performance of LLMs while using far less computation and energy. Restricting training to the balanced Cookie Theft subset and running all inference locally removed the privacy and regulatory issues common with cloud LLM deployments. The top SLM, DeepSeek R1 1.5 B, reached 91 % accuracy and F1 score, exceeded prior LLM baselines, and generated interpretable reasoning traces that can support clinical decisions. These findings indicate that smaller, fully fine-tuned models can streamline cognitive workflows and help clinicians make faster, more confident diagnostic decisions.

## Data Availability

All data produced in the present study are available upon reasonable request to the authors of the DementiaBank database.
All the data can be found online at: https://talkbank.org/dementia/access/English/Pitt.html

## Acknowledgements

The project has been supported by the Alzheimer’s Association Research Grant (24AARGD-NTF-1242722). There is no commercial interest to be disclosed.

## References

1. Alzheimer, Association. American Perspectives on Early Detection of Alzheimer’s Disease in the Era of Treatment.

2. Fang M, Hu J, Weiss J, et al. Lifetime risk and projected burden of dementia. Nat Med. 2025;31(3):772–776. doi:10.1038/S41591-024-03340-9

3. Cox CG, Brush BL, Kobayashi LC, Roberts JS. Determinants of dementia diagnosis in U.S. primary care in the past decade: A scoping review. J Prev Alzheimers Dis. 2025;12(2):100035. doi:10.1016/J.TJPAD.2024.100035

4. Dementia diagnosis, treatment delayed due to underuse of early detection tools, study finds - McKnight’s Senior Living. Accessed August 24, 2025. https://www.mcknightsseniorliving.com/news/dementia-diagnosis-treatment-delayed-due-to-underuse-of-early-detection-tools-study-finds/

5. PET MRI For Dementia Care | RSNA. Accessed September 14, 2025. https://www.rsna.org/news/2024/may/pet-mri-dementia-care

6. Mudrazija S, Aranda MP, Gaskin DJ, Monroe S, Richard P. Economic Burden of Alzheimer Disease and Related Dementias by Race and Ethnicity, 2020 to 2060. JAMA Netw Open. 2025;8(6):e2513931–e2513931. doi:10.1001/JAMANETWORKOPEN.2025.13931

7. Hansson O, Lehmann S, Otto M, Zetterberg H, Lewczuk P. Advantages and disadvantages of the use of the CSF Amyloid β (Aβ) 42/40 ratio in the diagnosis of Alzheimer’s Disease. Alzheimers Res Ther. 2019;11(1):34. doi:10.1186/S13195-019-0485-0

8. Hampel H, Hu Y, Cummings J, et al. Blood-based biomarkers for Alzheimer’s disease: Current state and future use in a transformed global healthcare landscape. Neuron. 2023;111(18):2781–2799. doi:10.1016/j.neuron.2023.05.017

9. Motter R, Vigo-Pelfrey C, Kholodenko D, et al. Reduction of β-amyloid peptide42 in the cerebrospinal fluid of patients with Alzheimer’s disease. Ann Neurol. 1995;38(4):643–648. doi:10.1002/ANA.410380413

10. Hansson O, Edelmayer RM, Boxer AL, et al. The Alzheimer’s Association appropriate use recommendations for blood biomarkers in Alzheimer’s disease. Alzheimer’s & Dementia. 2022;18(12):2669. doi:10.1002/ALZ.12756

11. Zhang Y, Wang Y, Shi C, Shen M, Lu F. Advances in retina imaging as potential biomarkers for early diagnosis of Alzheimer’s disease. Transl Neurodegener. 2021;10(1):1–9. doi:10.1186/S40035-021-00230-9/TABLES/2

12. Wang G, Estrella A, Hakim O, et al. Mini-Mental State Examination and Montreal Cognitive Assessment as Tools for Following Cognitive Changes in Alzheimer’s Disease Neuroimaging Initiative Participants. Journal of Alzheimer’s Disease. 2022;90(1):263–270. doi:10.3233/JAD-220397,

13. Beltrami D, Gagliardi G, Favretti RR, Ghidoni E, Tamburini F, Calzà L. Speech analysis by natural language processing techniques: A possible tool for very early detection of cognitive decline? Front Aging Neurosci. 2018;10:369. doi:10.3389/FNAGI.2018.00369/FULL

14. Orange JB, Molloy DW, Lever JA, Darzins P, Ganesan CR. Alzheimer’s disease. Physician-patient communication. Canadian Family Physician. 1994;40:1160. Accessed August 24, 2025. https://pmc.ncbi.nlm.nih.gov/articles/PMC2380218/

15. Chou CJ, Chang CT, Chang YN, et al. Screening for early Alzheimer’s disease: enhancing diagnosis with linguistic features and biomarkers. Front Aging Neurosci. 2024;16:1451326. doi:10.3389/FNAGI.2024.1451326/BIBTEX

16. Wang G, Estrella A, Hakim O, et al. Mini-Mental State Examination and Montreal Cognitive Assessment as Tools for Following Cognitive Changes in Alzheimer’s Disease Neuroimaging Initiative Participants. Journal of Alzheimer’s Disease. 2022;90(1):263–270. doi:10.3233/JAD-220397,

17. Johnson AEW, Pollard TJ, Shen L, et al. MIMIC-III, a freely accessible critical care database. Sci Data. 2016;3(1):1–9. doi:10.1038/SDATA.2016.35;SUBJMETA=139,1750,308,409,692,700;KWRD=DIAGNOSIS,HEALTH+CARE,MEDICAL+RESEARCH,OUTCOMES+RESEARCH,PROGNOSIS

18. Wong PC, Abdullah SS, Shapiai MI. Exceptional performance with minimal data using a generative adversarial network for alzheimer’s disease classification. Sci Rep. 2024;14(1):1–13. doi:10.1038/S41598-024-66874-5;SUBJMETA=1283,166,365,375,639,692,699,987;KWRD=ALZHEIMER

19. Diogo VS, Ferreira HA, Prata D. Early diagnosis of Alzheimer’s disease using machine learning: a multi-diagnostic, generalizable approach. Alzheimers Res Ther. 2022;14(1):107. doi:10.1186/S13195-022-01047-Y

20. Mohsen S. Alzheimer’s disease detection using deep learning and machine learning: a review. Artif Intell Rev. 2025;58(9):1–39. doi:10.1007/S10462-025-11258-Y/TABLES/8

21. Malik MM. A Hierarchy of Limitations in Machine Learning. Published online 2020. Accessed September 8, 2025. https://arxiv.org/abs/2002.05193.

22. Deep learning vs. machine learning - Azure Machine Learning | Microsoft Learn. Accessed September 9, 2025. https://learn.microsoft.com/en-us/azure/machine-learning/concept-deep-learning-vs-machine-learning?view=azureml-api-2

23. El-Latif AAA, Chelloug SA, Alabdulhafith M, Hammad M. Accurate Detection of Alzheimer’s Disease Using Lightweight Deep Learning Model on MRI Data. Diagnostics. 2023;13(7):1216. doi:10.3390/DIAGNOSTICS13071216

24. Ksibi A, Walha A, Zakariah M, Ayadi M, Alshalali T, Almujally NA. Multimodal Siamese networks for dementia detection from speech in women. Sci Rep. 2025;15(1):1–32. doi:10.1038/S41598-025-13902-7;SUBJMETA

25. Nawaz A, Iqbal J, Anwar SM, Liaqat R, Majid M. Deep Convolutional Neural Network based Classification of Alzheimer’s Disease using MRI Data. Accessed September 14, 2025. http://www.loni.ucla.edu/ADNI

26. Aramaki E, Wakamiya S, Yada S, Nakamura Y. Natural Language Processing: from Bedside to Everywhere. Yearb Med Inform. 2022;31(1):243. doi:10.1055/S-0042-1742510

27. Karanikolas N, Manga E, Samaridi N, Tousidou E, Vassilakopoulos M. Large Language Models versus Natural Language Understanding and Generation. ACM International Conference Proceeding Series. Published online November 24, 2023:278–290. doi:10.1145/3635059.3635104

28. Dennstädt F, Hastings J, Putora PM, Schmerder M, Cihoric N. Implementing large language models in healthcare while balancing control, collaboration, costs and security. NPJ Digit Med. 2025;8(1):143. doi:10.1038/S41746-025-01476-7

29. Shanmugarasa Y, Ding M, Arachchige CM, Rakotoarivelo T. SoK: The Privacy Paradox of Large Language Models: Advancements, Privacy Risks, and Mitigation. Proceedings of the 20th ACM Asia Conference on Computer and Communications Security. 2025;1:425–441. doi:10.1145/3708821.3733888

30. Comparing Cloud-Based vs Local Deployment of Large Language Models (LLMs): Advantages and Disadvantages - CertLibrary Blog. Accessed September 9, 2025. https://www.certlibrary.com/blog/comparing-cloud-based-vs-local-deployment-of-large-language-models-llms-advantages-and-disadvantages/

31. Belcak P, Heinrich G, Diao S, et al. Small Language Models are the Future of Agentic AI. Published online 2025.

32. DementiaBank English Pitt Corpus. Accessed September 25, 2025. https://talkbank.org/dementia/access/English/Pitt.html

33. Spreen Otfried, Risser AH. Assessment of aphasia. Published online 2003:320.

34. Using CLAN. TalkBank. Accessed September 25, 2025. http://dali.talkbank.org/clan/

35. Wei J, Wang X, Schuurmans D, et al. Chain-of-Thought Prompting Elicits Reasoning in Large Language Models. Adv Neural Inf Process Syst. 2022;35. Accessed September 25, 2025. https://arxiv.org/pdf/2201.11903

36. Van Nguyen C, Shen X, Aponte R, et al. A Survey of Small Language Models. Published online October 25, 2024. Accessed September 16, 2025. https://arxiv.org/pdf/2410.20011

37. Advantages of Small Language Models Over Large Language Models? | by Eastgate Software | Medium. Accessed September 23, 2025. https://medium.com/@eastgate/advantages-of-small-language-models-over-large-language-models-a52deb47d50b

38. DeepSeek-AI. DeepSeek-R1. Published online 2025. Accessed September 25, 2025. https://arxiv.org/pdf/2501.12948v1

39. Guo D, Zhu Q, Yang D, et al. DeepSeek-Coder: When the Large Language Model Meets Programming-The Rise of Code Intelligence. Published online 2024. Accessed August 25, 2025. https://github.com/deepseek-ai/DeepSeek-Coder

40. Chaudhary Pr M, Geiger Pr A, Group R. Evaluating Open-Source Sparse Autoencoders on Disentangling Factual Knowledge in GPT-2 Small.

41. Grattafiori A, Dubey A, Jauhri A, et al. The Llama 3 Herd of Models. Published online July 31, 2024. Accessed July 17, 2025. https://arxiv.org/pdf/2407.21783

42. Mo T, K Lam JC, K Li VO, L Cheung LY. Leveraging Large Language Models for Identifying Interpretable Linguistic Markers and Enhancing Alzheimer’s Disease. doi:10.1101/2024.08.22.24312463

43. Zheng T, Xie X, Peng X, Chen H, Tian F. Alzheimer’s disease detection based on large language model prompt engineering.

44. Lin K, Washington PY. Multimodal deep learning for dementia classification using text and audio. Sci Rep. 2024;14(1):1–10. doi:10.1038/S41598-024-64438-1;SUBJMETA=114,2397,631,692,700;KWRD=COMPUTATIONAL+MODELS,HEALTH+CARE

45. Mehdoui SS, Bouzid A, Sierra-Sosa D, Elmaghraby A. Dementia Insights: A Context-Based MultiModal Approach.

46. Lei Y. Training strategies, computational consumption, and memory control for large-scale language models. Proceedings of the 2025 5th International Conference on Applied Mathematics, Modelling and Intelligent Computing, CAMMIC 2025. Published online August 25, 2025:786–790. doi:10.1145/3745533.3745660;SUBPAGE:STRING:FULL

47. Busch F, Hoffmann L, Rueger C, et al. Current applications and challenges in large language models for patient care: a systematic review. Communications Medicine 2025 5:1. 2025;5(1):26-. doi:10.1038/s43856-024-00717-2

48. Artsi Y, Sorin V, Glicksberg BS, et al. Challenges of Implementing LLMs in Clinical Practice: Perspectives. J Clin Med. 2025;14(17):6169. doi:10.3390/JCM14176169

49. Alber DA, Yang Z, Alyakin A, et al. Medical large language models are vulnerable to data-poisoning attacks. Nat Med. 2025;31(2):618. doi:10.1038/S41591-024-03445-1

50. The (not so) Hidden Risks of Using Cloud-Based LLMs with Confidential Data – ALPHALECT.ai. Accessed November 29, 2025. https://alphalect.ai/blog/the-not-so-hidden-risks-of-using-cloud%E2%80%91based-llms-with-confidential-data/

51. Christof M, Armoundas AA. Implications of integrating large language models into clinical decision making. Communications Medicine 2025 5:1. 2025;5(1):490-. doi:10.1038/s43856-025-01216-8

52. Shojaee P, Mirzadeh I, Alizadeh K, Horton M, Bengio S, Farajtabar M. The illusion of thinking: Understanding the strengths and limitations of reasoning models via the lens of problem complexity. arXiv preprint arXiv:250606941. Published online 2025.

53. Pool J, Indulska M, Sadiq S. Large language models and generative AI in telehealth: a responsible use lens. Published online 2024. doi:10.1093/jamia/ocae035

